# Program and patient characteristics for the United States Expanded Access Program to COVID-19 convalescent plasma

**DOI:** 10.1101/2021.04.08.21255115

**Authors:** Jonathon W. Senefeld, Patrick W. Johnson, Katie L. Kunze, Noud van Helmond, Stephen A. Klassen, Chad C. Wiggins, Katelyn A. Bruno, Michael A. Golafshar, Molly M. Petersen, Matthew R. Buras, Allan M. Klompas, Matthew A. Sexton, Juan C. Diaz Soto, Sarah E. Baker, John R.A. Shepherd, Nicole C. Verdun, Peter Marks, Camille M. van Buskirk, Jeffrey L. Winters, James R. Stubbs, Robert F. Rea, Vitaly Herasevich, Emily R. Whelan, Andrew J. Clayburn, Kathryn F. Larson, Juan G. Ripoll, Kylie J. Andersen, Matthew N.P. Vogt, Joshua J. Dennis, Riley J. Regimbal, Philippe R. Bauer, Janis E. Blair, Katherine Wright, Joel T. Greenshields, Nigel S. Paneth, DeLisa Fairweather, R. Scott Wright, Arturo Casadevall, Rickey E. Carter, Michael J. Joyner

## Abstract

**Background:** The United States (US) Expanded Access Program (EAP) to COVID-19 convalescent plasma was initiated in response to the rapid spread of severe acute respiratory syndrome coronavirus 2 (SARS-CoV-2), the causative agent of coronavirus disease-2019 (COVID-19). While randomized clinical trials were in various stages of development and enrollment, there was an urgent need for widespread access to potential therapeutic agents particularly for vulnerable racial and ethnic minority populations who were disproportionately affected by the pandemic. The objective of this study is to report on the demographic, geographic, and chronological access to COVID-19 convalescent plasma in the US via the EAP.

**Methods and findings:** Mayo Clinic served as the central IRB for all participating facilities and any US physician could participate as local physician–principal investigator. Registration occurred through the EAP central website. Blood banks rapidly developed logistics to provide convalescent plasma to hospitalized patients with COVID-19. Demographic and clinical characteristics of all enrolled patients in the EAP were summarized. Temporal trends in access to COVID-19 convalescent plasma were investigated by comparing daily and weekly changes in EAP enrollment in response to changes in infection rate on a state level. Geographical analyses on access to convalescent plasma included assessing EAP enrollment in all national hospital referral regions as well as assessing enrollment in metropolitan and less populated areas which did not have access to COVID-19 clinical trials.

From April 3 to August 23, 2020, 105,717 hospitalized patients with severe or life-threatening COVID-19 were enrolled in the EAP. A majority of patients were older than 60 years of age (57.8%), male (58.4%), and overweight or obese (83.8%). There was substantial inclusion of minorities and underserved populations, including 46.4% of patients with a race other than White, and 37.2% of patients were of Hispanic ethnicity. Severe or life-threatening COVID-19 was present in 61.8% of patients and 18.9% of patients were mechanically ventilated at time of convalescent plasma infusion. Chronologically and geographically, increases in enrollment in the EAP closely followed confirmed infections across all 50 states. Nearly all national hospital referral regions enrolled patients in the EAP, including both in metropolitan and less populated areas.

**Conclusions:** The EAP successfully provided widespread access to COVID-19 convalescent plasma in all 50 states, including for underserved racial and ethnic minority populations. The efficient study design of the EAP may serve as an example framework for future efforts when broad access to a treatment is needed in response to a dynamic disease affecting demographic groups and areas historically underrepresented in clinical studies.

## Introduction

Severe acute respiratory syndrome coronavirus 2 (SARS-CoV-2), the causative agent of the coronavirus disease 2019 (COVID-19), spread rapidly across the United States (US) after confirmation of the first case of COVID-19 in the US in December 2019 and January 2020 [1]. By March of 2020 in the Northeast region of the US, community transmission was occurring in major metropolitan areas and hospitals became overwhelmed with admissions for severe or life-threating COVID-19 [1]. Although most infected persons have few or no symptoms despite high SARS-CoV-2 viral loads [2], a smaller group of persons develop hypoxemia and severe COVID-19 leading to hospitalization with supplemental oxygen support [3]. Severe cases of COVID-19 can lead to respiratory failure, which is among the leading causes of death in persons with COVID-19 [4].

The cornerstone of treatment for patients with COVID-19 was symptomatic and supportive management [5]. Because there was a dearth of evidence supporting the efficacy of COVID-19 therapeutic strategies during the early stages of this public health emergency, immunomodulatory agents and antivirals, represented promising strategies that were anticipated to provide benefit to patients with COVID-19 [6, 7]. Passive immunotherapy using convalescent plasma or serum had been used previously to treat acute respiratory tract infections [8-10], including severe acute respiratory syndrome coronavirus 1 infection (SARS-CoV-1) [11], and early studies suggested potential efficacy of convalescent plasma in the treatment of COVID-19 [12-14].

In late March and early April of 2020, COVID-19 convalescent plasma began to be administered to patients under single-patient emergency investigational new drug (eIND) applications while randomized clinical trials of COVID-19 convalescent plasma were in various stages of development and enrollment. For example, one institution in New York City had 45 eIND applications submitted to and approved by the US Food and Drug Administration (FDA) in a two-week period in late March 2020 [15]. The single-patient eIND application process requires substantial administrative support from local institutions and the US FDA and limits widespread and high-volume access to convalescent plasma, particularly for underserved and racial and ethnic minority populations, who were disproportionately affected by the COVID-19 pandemic [16, 17]. Additionally, clinical trials often have inclusion criteria that are restricted to a specific geographical region or disease status (e.g. hospitalized, but not severe patients) and have exclusion criteria (e.g. prisoners or recipients of solid organ transplant). Thus, inherently, more traditional regulatory pathways for obtaining access to COVID-19 convalescent plasma (eIND or clinical trials) result in limited access to convalescent plasma and may potentially limit the ability to comprehensively study the safety of convalescent plasma for the treatment of COVID-19.

To provide access to a COVID-19 treatment and provide a framework for standardized safety data collection, Mayo Clinic initiated the Expanded Access Program (EAP) for COVID-19 convalescent plasma. The primary objective of the EAP was to provide access to convalescent plasma for hospitalized patients in the US with severe or life-threatening COVID-19 [18]. The EAP started as a national registry approved to enroll 5,000 patients, but due to a national demand for COVID-19 plasma, enrollment goals were extended in collaboration with the US Food and Drug Administration (FDA) and the Biomedical Advanced Research and Development Authority (BARDA) with the aim for the EAP to become a broad national program obviating the need for individual patient INDs. We herein assess the extent to which the EAP was successful in terms of providing access to COVID-19 convalescent plasma by presenting demographic, geographic, and chronological characteristics of enrollment in the EAP alongside publicly available data of state-level trends in COVID-19.

## Methods

As described previously [19-21], the EAP was a national registry for hospitalized patients with COVID-19. Collaborative support was provided by the US BARDA and FDA; funding to support the study infrastructure and study-related costs at participating sites was provided under contract from BARDA. Mayo Clinic served as the academic research organization coordinating the national registry. The Mayo Clinic Institutional Review Board (IRB), the central IRB for the registry, approved the protocol (IRB #20-0033412, NCT#: NCT04338360), all amendments, and provided regulatory oversight for all sites and investigators. The principal investigator (M.J.J.) was the regulatory sponsor. A Data and Safety Monitoring Board oversaw the safety analyses and advised the regulatory sponsor and the Mayo Clinic IRB on risk.

### Patients

Patients were eligible for enrollment in the EAP if they were: aged 18 years or older, hospitalized with a laboratory confirmed diagnosis of or suspected/probable infection with severe acute respiratory syndrome coronavirus 2 (SARS-CoV-2), and either had or were judged by a healthcare provider to be at high risk of progression to severe or life-threatening COVID-19. Severe COVID-19 was defined by one or more of the following: dyspnea, respiratory frequency ≥ 30/min, blood oxygen saturation ≤ 93%, partial pressure of arterial oxygen to fraction of inspired oxygen ratio < 300, lung infiltrates > 50% within 24 to 48 hours of hospital admission. Life-threatening COVID-19 was defined as one or more of the following: respiratory failure, septic shock, and multiple organ dysfunction or failure. To maximize access to COVID-19 convalescent plasma, no exclusion criteria were used, thereby, enabling access to vulnerable adult populations who may not be eligible for clinical trials, including pregnant women and prisoners.

### Enrollment

All hospitals and acute care facilities in the US and its territories, and any physician licensed in the US, were allowed to register for participation provided they agreed to adhere to the online available treatment protocol [18], as well as US FDA and state regulations. All patient registration was facilitated through the central study website [22]. A single consent form, available in eight languages, was used by all participating sites. Informed consent was obtained from the patient or a legally authorized representative prior to enrollment, except for patients in whom an emergency consent was utilized. Criteria for emergency consent were consistent with the federal regulation governing emergency consent [23]. COVID-19 convalescent plasma transfusate and convalescent plasma donors details have been described elsewhere [19-21].

### Study data

Demographic and clinical characteristics of enrolled patients were collected using the Research Electronic Data Capture system (REDCap, v.9.1.15 – v10.0.33 Vanderbilt University, Nashville, TN) [24, 25]. REDCap is US Health Insurance Portability and Accountability Act (HIPAA) compliant and the US FDA authorized its use in the EAP [20, 21]. The EAP was expeditiously established and implemented to provide access to a potential therapeutic for COVID-19 at a time when no satisfactory therapies for COVID-19 were available. To maximize access in the context of stress on clinical staff at participating sites during the COVID-19 pandemic, the online case reporting forms were designed to optimize convenience. Race (American Indian or Alaska Native, Asian, Black or African American, Native Hawaiian or Other Pacific Islander, White) and ethnicity (Hispanic or Latino, Not Hispanic or Latino) were reported into categories by site personnel consistent with guidelines provided by the US Office of Management and Budget [26].

The database was updated as needed to fulfill the requirements of the EAP IRB and data collection requirements of BARDA. As the original goal of data collection was to determine safety, updates were needed to capture additional data as enrollment expanded and the study progressed. Enrollment into the EAP was stopped after the FDA issued an emergency use authorization (EUA) for COVID-19 convalescent plasma on August 23rd, 2020. Data clarification requests were sent to participating investigators as needed until the database was locked to further data changes on December 16th, 2020. The study protocol, case report forms with completion instructions, and informed consent form are publicly available on the study website [22], archived by the US Library of Congress.

### COVID-19 epidemiological data sources

In order to contextualize whether the patients enrolled in the EAP were reflective of the US population, race and ethnicity data of each state and US territories were retrieved from the US Census Bureau [27], using the same race and ethnic categories that were collected in the EAP. COVID-19-confirmed infection rates per day for each US state were obtained from the New York Times database [28]. Hospital referral regions are regional healthcare markets defined by where the majority of residents within that region have their hospitalization stays. The 306 hospital referral regions in the US were retrieved from the Dartmouth Atlas of Health Care. Metropolitan and less populated US region data were obtained from US Census Bureau data [27] and the 2010 Office of Management and Budget Standards that define metropolitan and micropolitan areas based on statistical assessments [29]. Micropolitan areas were defined as areas with a population of at least 10,000 and less than 50,000 residents. The US regions of Northeast, Southeast, Midwest, Southwest, and West were delineated using commonly used regions [30]. Characteristics of US hospitals were retrieved from American Hospital Directory [31] and the Centers for Medicare and Medicaid [32]. The potential limitations of the data sources are described in the discussion.

### Statistical Considerations

To provide a comprehensive report of the enrollment data on the EAP program descriptive statistics are presented for demographic and clinical variables of interest. To examine enrollment in the EAP over time, dot plots were used to show the number of enrollments, for each US state individually and aggregated by region by day of the study. Additionally, EAP enrollment was compared to the number of confirmed COVID-19 cases per state over the duration of the study. During the window of EAP enrollment, a moving 7-day average was calculated for daily enrollments and COVID-19 cases within each state that enrolled more than 10 patients in total in the EAP. To compare and visualize relative patterns, these averages were scaled between 0 (lowest cases/enrollments) and 1 (peak cases/enrollments) and overlaid on a geo-faceted graph. The geo-faceted graph contains one cell for each US state and is placed at approximately the same location on the graph as the corresponding geographical location. Differences in geographic access to convalescent plasma through the EAP were assessed by examining enrollment across micropolitan and metropolitan areas and the number of hospitals enrolling patients in each hospital referral region in the United States. All data were processed using R version 3.6.2.

## Results

### Patient Characteristics

From April 3rd to August 23rd, 2020, 105,717 hospitalized patients with severe or life-threatening COVID-19 were enrolled in the EAP and ∼95,000 patients were transfused with COVID-19 convalescent plasma. The EAP halted enrollment forthwith after the US FDA issued an emergency use authorization (EUA) for COVID-19 convalescent plasma, citing that the totality of scientific evidence indicated that convalescent plasma was safe [20, 21] and a potentially promising therapeutic treatment on August 23, 2020 [33]. This authorization enabled physicians to use COVID-19 convalescent plasma without requesting IND permission and obviated the need for access to convalescent plasma via the EAP.

Enrolled patients’ demographic characteristics including age, gender, race, ethnicity, and other clinical variables at time of transfusion are shown in **Table 1**. A majority of patients were older than 60 years of age (57.8%), male (58.4%), overweight or obese (83.8%), and had never smoked (69.7%). There was substantial inclusion of minorities and underserved populations, including 46.4% of patients with a race other than White, and 37.2% of patients were of Hispanic ethnicity. Pre-existing conditions present among enrolled patients are displayed in **Supplemental Table 1**. Of those patients enrolled, 61.8% had severe or life-threatening COVID-19, 42.3% were in the ICU, and 18.9% received mechanical ventilation at time of transfusion. A small proportion of patients (3.9%) had no form of hospital respiratory support prior to infusion. A large percentage of patients had severe risk factors of dyspnea (75.7%), oxygen saturation ≤ 93% (75.0%), and acute respiratory failure (60.6%). Patients were also prescribed steroids (65.7%), azithromycin (49.0%) and remdesivir (37.6%) during their hospital stay. The median number of days between diagnosis of COVID-19 and the first transfusion was 4 days (interquartile range, 2 – 8 days), and nearly half of transfused patients (45.0%) received convalescent plasma within 3 days of COVID-19 diagnosis, which often occurred during hospital admission.

**Table 1.**
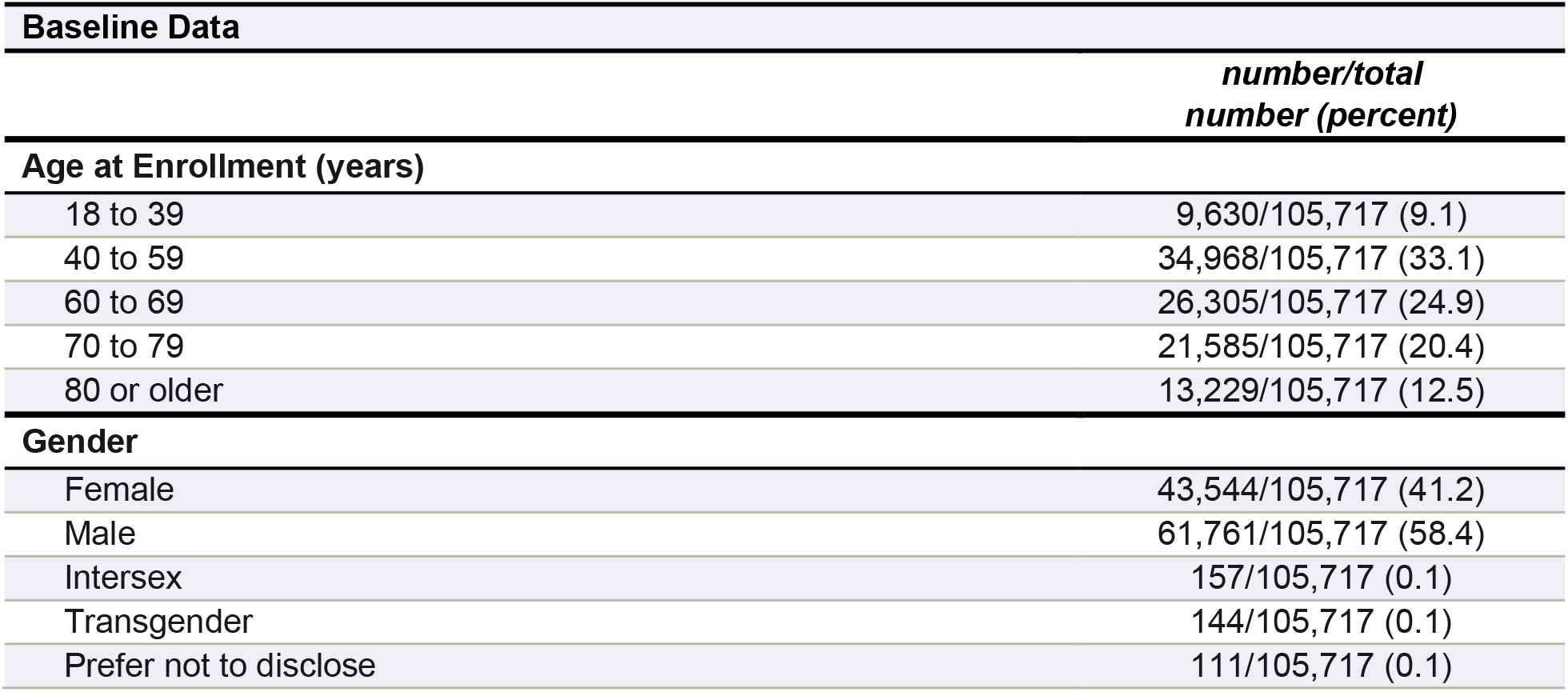
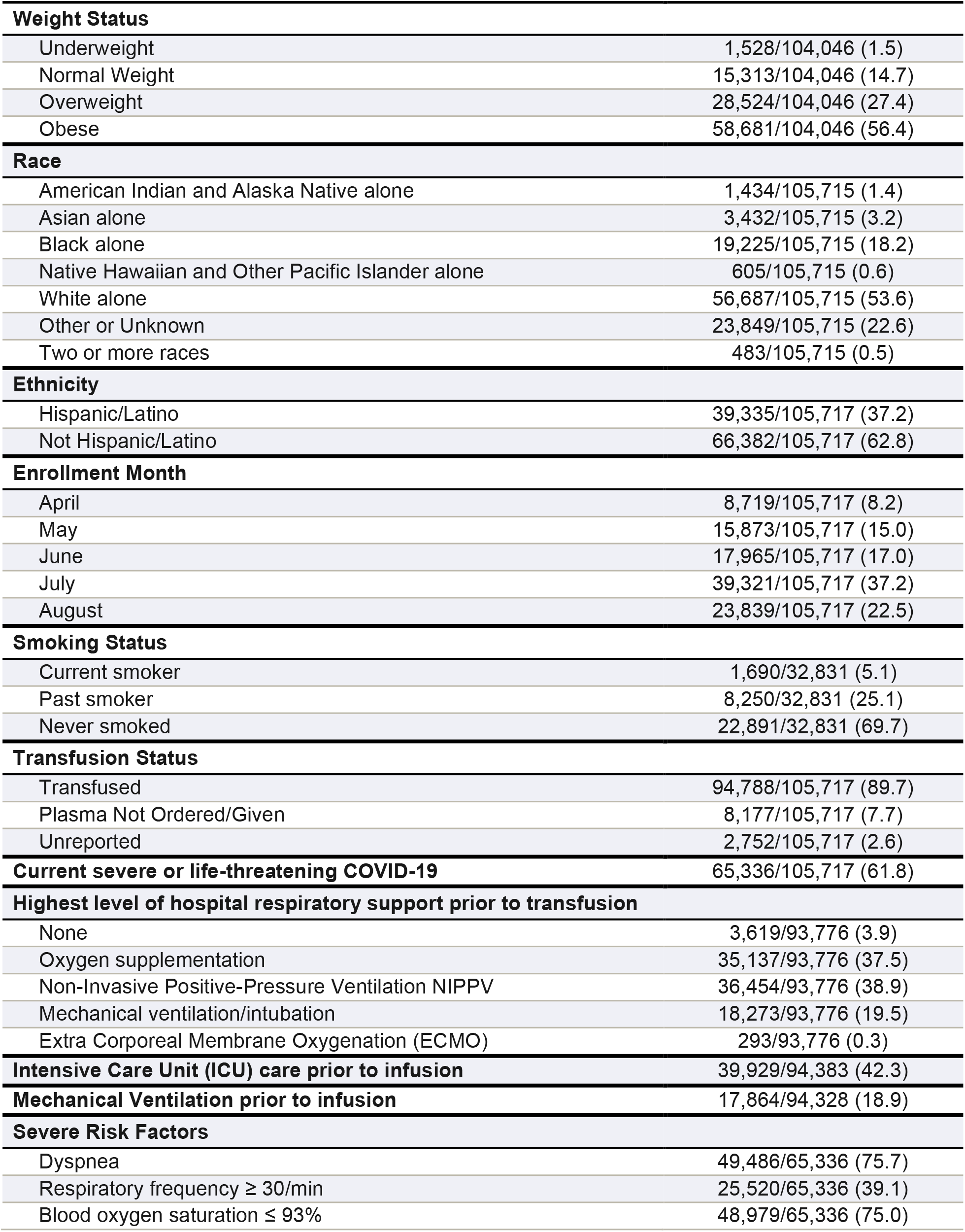
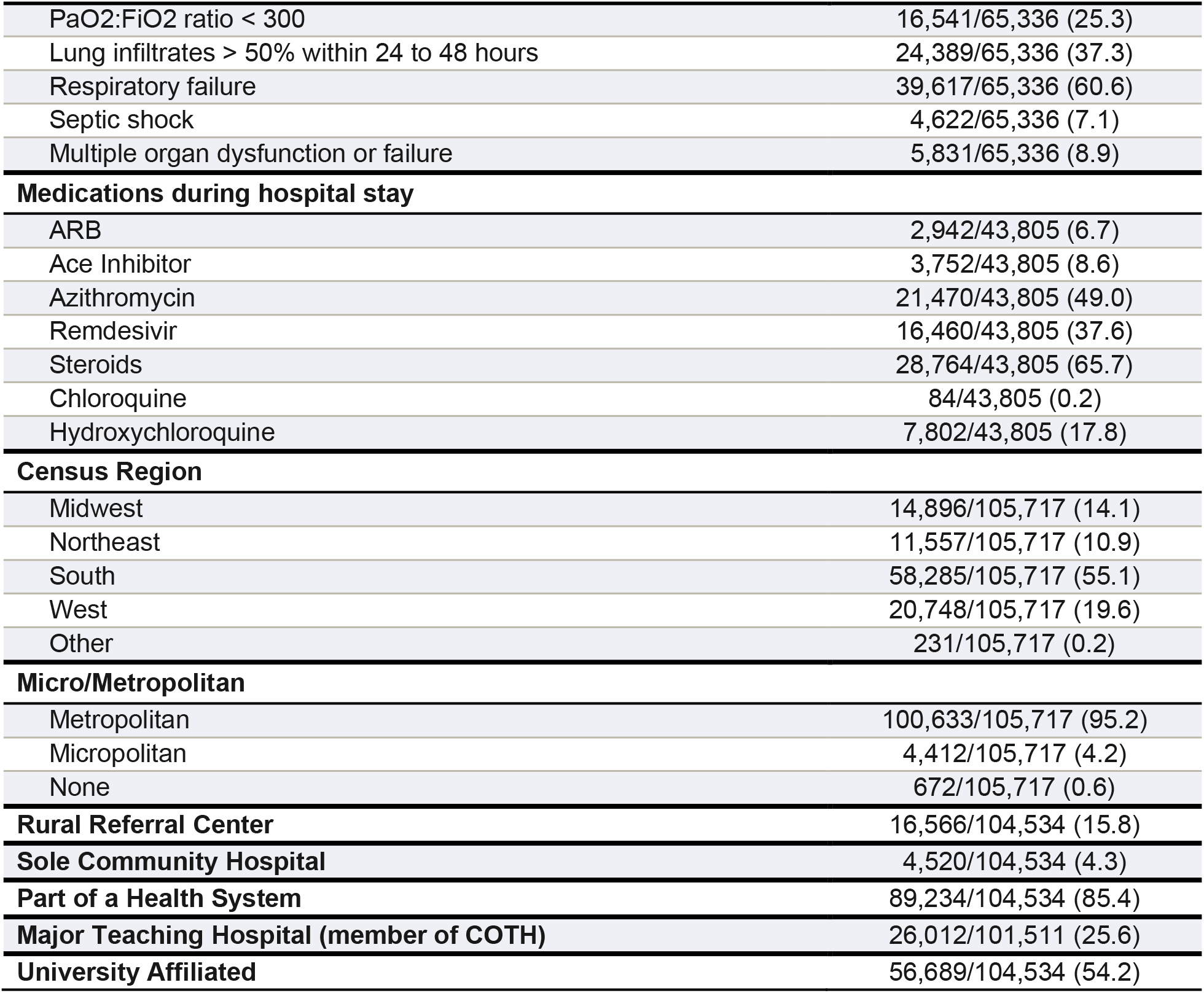
Characteristics of Patients with COVID-19 who enrolled in the US Expanded Access Program to convalescent plasma.

| **Baseline Data** |  |
| --- | --- |
|  | *number/total*<br>*number (percent)* |
| **Age at Enrollment (years)** |  |
| 18 to 39 | 9,630/105,717 (9.1) |
| 40 to 59 | 34,968/105,717 (33.1) |
| 60 to 69 | 26,305/105,717 (24.9) |
| 70 to 79 | 21,585/105,717 (20.4) |
| 80 or older | 13,229/105,717 (12.5) |
| **Gender** |  |
| Female | 43,544/105,717 (41.2) |
| Male | 61,761/105,717 (58.4) |
| Intersex | 157/105,717 (0.1) |
| Transgender | 144/105,717 (0.1) |
| Prefer not to disclose | 111/105,717 (0.1) |
|  | *number/total*<br>*number (percent)* |
| **Weight Status** |  |
| Underweight | 1,528/104,046 (1.5) |
| Normal Weight | 15,313/104,046 (14.7) |
| Overweight | 28,524/104,046 (27.4) |
| Obese | 58,681/104,046 (56.4) |
| **Race** |  |
| American Indian and Alaska Native alone | 1,434/105,715 (1.4) |
| Asian alone | 3,432/105,715 (3.2) |
| Black alone | 19,225/105,715 (18.2) |
| Native Hawaiian and Other Pacific Islander alone | 605/105,715 (0.6) |
| White alone | 56,687/105,715 (53.6) |
| Other or Unknown | 23,849/105,715 (22.6) |
| Two or more races | 483/105,715 (0.5) |
| **Ethnicity** |  |
| Hispanic/Latino | 39,335/105,717 (37.2) |
| Not Hispanic/Latino | 66,382/105,717 (62.8) |
| **Enrollment Month** |  |
| April | 8,719/105,717 (8.2) |
| May | 15,873/105,717 (15.0) |
| June | 17,965/105,717 (17.0) |
| July | 39,321/105,717 (37.2) |
| August | 23,839/105,717 (22.5) |
| **Smoking Status** |  |
| Current smoker | 1,690/32,831 (5.1) |
| Past smoker | 8,250/32,831 (25.1) |
| Never smoked | 22,891/32,831 (69.7) |
| **Transfusion Status** |  |
| Transfused | 94,788/105,717 (89.7) |
| Plasma Not Ordered/Given | 8,177/105,717 (7.7) |
| Unreported | 2,752/105,717 (2.6) |
| **Current severe or life-threatening COVID-19** | 65,336/105,717 (61.8) |
| **Highest level of hospital respiratory support prior to transfusion** |  |
| None | 3,619/93,776 (3.9) |
| Oxygen supplementation | 35,137/93,776 (37.5) |
| Non-Invasive Positive-Pressure Ventilation NIPPV | 36,454/93,776 (38.9) |
| Mechanical ventilation/intubation | 18,273/93,776 (19.5) |
| Extra Corporeal Membrane Oxygenation (ECMO) | 293/93,776 (0.3) |
| **Intensive Care Unit (ICU) care prior to infusion** | 39,929/94,383 (42.3) |
| **Mechanical Ventilation prior to infusion** | 17,864/94,328 (18.9) |
| **Severe Risk Factors** |  |
| Dyspnea | 49,486/65,336 (75.7) |
| Respiratory frequency ≥ 30/min | 25,520/65,336 (39.1) |
| Blood oxygen saturation ≤ 93% | 48,979/65,336 (75.0) |
|  | *number/total*<br>*number (percent)* |
| PaO<sub>2</sub>:FiO<sub>2</sub> ratio < 300 | 16,541/65,336 (25.3) |
| Lung infiltrates > 50% within 24 to 48 hours | 24,389/65,336 (37.3) |
| Respiratory failure | 39,617/65,336 (60.6) |
| Septic shock | 4,622/65,336 (7.1) |
| Multiple organ dysfunction or failure | 5,831/65,336 (8.9) |
| **Medications during hospital stay** |  |
| ARB | 2,942/43,805 (6.7) |
| Ace Inhibitor | 3,752/43,805 (8.6) |
| Azithromycin | 21,470/43,805 (49.0) |
| Remdesivir | 16,460/43,805 (37.6) |
| Steroids | 28,764/43,805 (65.7) |
| Chloroquine | 84/43,805 (0.2) |
| Hydroxychloroquine | 7,802/43,805 (17.8) |
| **Census Region** |  |
| Midwest | 14,896/105,717 (14.1) |
| Northeast | 11,557/105,717 (10.9) |
| South | 58,285/105,717 (55.1) |
| West | 20,748/105,717 (19.6) |
| Other | 231/105,717 (0.2) |
| **Micro/Metropolitan** |  |
| Metropolitan | 100,633/105,717 (95.2) |
| Micropolitan | 4,412/105,717 (4.2) |
| None | 672/105,717 (0.6) |
| **Rural Referral Center** | 16,566/104,534 (15.8) |
| **Sole Community Hospital** | 4,520/104,534 (4.3) |
| **Part of a Health System** | 89,234/104,534 (85.4) |
| **Major Teaching Hospital (member of COTH)** | 26,012/101,511 (25.6) |
| **University Affiliated** | 56,689/104,534 (54.2) |

### Geospatial Trends in Enrollment

Patients were enrolled from each state in the US, the District of Columbia, and the US territories of Puerto Rico, and the US Virgin Islands (**Table 2**). A large percentage of patients were enrolled in the Southern region of the US (55.1%) and most patients enrolled in a hospital within a metropolitan area (95.2%) that was part of a health system (85.4%) and/or was university affiliated (54.2%).

**Table 2.** Tabular summaries of patient enrollment in the US Expanded Access Program (EAP) to convalescent plasma and US COVID-19 cases during the EAP enrollment period, stratified by US state or territory in alphabetical order.

| State | EAP Summaries |  |  | Population and COVID+ Ratios |  |  |
| --- | --- | --- | --- | --- | --- | --- |
|  | Enrolling Sites | Enrolled Patients | Transfused Patients | COVID+ Cases / Enrollment | Enrollments / 10,000 people | Enrollments / 1,000 COVID+ Cases |
| Alabama | 35 | 2,408 | 2,016 | 51.9 | 4.9 | 19.3 |
| Alaska | 3 | 59 | 58 | 100.5 | 0.8 | 9.9 |
| Arizona | 41 | 3,961 | 3,680 | 50.6 | 5.4 | 19.8 |
| Arkansas | 16 | 597 | 498 | 101.5 | 2.0 | 9.9 |
| California | 236 | 11,874 | 10,076 | 59.2 | 3.0 | 16.9 |
| Colorado | 27 | 646 | 628 | 84.3 | 1.1 | 11.9 |
| Connecticut | 23 | 1,022 | 928 | 48.3 | 2.9 | 20.7 |
| Delaware | 4 | 372 | 366 | 45.9 | 3.8 | 21.8 |
| District of Columbia | 9 | 349 | 278 | 38.4 | 4.9 | 26.0 |
| Florida | 158 | 12,553 | 11,206 | 49.0 | 5.8 | 20.4 |
| Georgia | 74 | 5,424 | 4,921 | 46.0 | 5.1 | 21.7 |
| Hawaii | 8 | 320 | 316 | 25.6 | 2.3 | 39.1 |
| Idaho | 10 | 156 | 131 | 202.5 | 0.9 | 4.9 |
| Illinois | 92 | 3,137 | 2,709 | 73.5 | 2.5 | 13.6 |
| Indiana | 64 | 1,746 | 1,610 | 53.7 | 2.6 | 18.6 |
| Iowa | 35 | 1,299 | 1,222 | 49.8 | 4.1 | 20.1 |
| Kansas | 23 | 703 | 609 | 61.0 | 2.4 | 16.4 |
| Kentucky | 40 | 884 | 836 | 56.8 | 2.0 | 17.6 |
| Louisiana | 44 | 1,548 | 1,351 | 92.0 | 3.3 | 10.9 |
| Maine | 9 | 62 | 58 | 67.5 | 0.5 | 14.8 |
| Maryland | 42 | 1,812 | 1,592 | 58.9 | 3.0 | 17.0 |
| Massachusetts | 37 | 907 | 790 | 133.2 | 1.3 | 7.5 |
| Michigan | 61 | 1,071 | 875 | 97.0 | 1.1 | 10.3 |
| Minnesota | 35 | 1,052 | 1,008 | 71.5 | 1.9 | 14.0 |
| Mississippi | 19 | 1,380 | 1,313 | 59.3 | 4.6 | 16.9 |
| Missouri | 41 | 1,347 | 1,262 | 62.9 | 2.2 | 15.9 |
| Montana | 8 | 133 | 127 | 54.3 | 1.2 | 18.4 |
| Nebraska | 18 | 509 | 471 | 66.9 | 2.6 | 15.0 |
| Nevada | 19 | 1,303 | 1,128 | 52.2 | 4.2 | 19.2 |
| New Hampshire | 8 | 127 | 92 | 54.0 | 0.9 | 18.5 |
| New Jersey | 66 | 2,768 | 2,452 | 62.0 | 3.1 | 16.1 |
| New Mexico | 14 | 433 | 415 | 57.7 | 2.1 | 17.3 |
| New York | 110 | 4,492 | 4,146 | 79.1 | 2.3 | 12.6 |
| North Carolina | 56 | 1,769 | 1,627 | 94.1 | 1.7 | 10.6 |
| North Dakota | 7 | 226 | 216 | 51.7 | 3.0 | 19.4 |
| Ohio | 77 | 2,184 | 2,007 | 55.2 | 1.9 | 18.1 |
| Oklahoma | 27 | 1,415 | 1,324 | 41.0 | 3.6 | 24.4 |
| Oregon | 17 | 320 | 290 | 81.2 | 0.8 | 12.3 |

| EAP Summaries |  |  |  | Population and COVID+ Ratios |  |  |
| --- | --- | --- | --- | --- | --- | --- |
| State | Enrolling Sites | Enrolled Patients | Transfused Patients | COVID+ Cases / Enrollment | Enrollments / 10,000 people | Enrollments / 1,000 COVID+ Cases |
| Pennsylvania | 90 | 1,930 | 1,702 | 68.9 | 1.5 | 14.5 |
| Puerto Rico | 31 | 222 | 184 | 148.3 | 0.7 | 6.7 |
| Rhode Island | 9 | 248 | 242 | 86.2 | 2.3 | 11.6 |
| South Carolina | 38 | 3,970 | 3,530 | 29.6 | 7.7 | 33.7 |
| South Dakota | 6 | 427 | 409 | 31.3 | 4.8 | 31.9 |
| Tennessee | 51 | 2,592 | 2,444 | 57.6 | 3.8 | 17.4 |
| Texas | 221 | 19,363 | 17,507 | 32.8 | 6.7 | 30.4 |
| Virgin Islands | 2 | 9 | 6 | 123.2 |  | 8.1 |
| Utah | 8 | 592 | 573 | 86.4 | 1.8 | 11.6 |
| Vermont | 1 | 1 | 1 | 1,303.0 | 0.0 | 0.8 |
| Virginia | 51 | 1,747 | 1,480 | 68.2 | 2.0 | 14.7 |
| Washington | 34 | 942 | 881 | 76.6 | 1.2 | 13.1 |
| West Virginia | 11 | 102 | 88 | 98.6 | 0.6 | 10.1 |
| Wisconsin | 62 | 1,195 | 1,100 | 66.3 | 2.1 | 15.1 |
| Wyoming | 4 | 9 | 9 | 411.6 | 0.2 | 2.4 |

Patients were enrolled by 12,879 healthcare providers from 2,722 hospitals and acute care facilities across the US, **Figure 1**. COVID-19 convalescent plasma was provided by 317 blood banks. The median number of participants per site was 22 (range, 1 to 1,175). While 2,722 sites were registered, 490 (18.0%) enrolled no patients, 732 (26.9%) enrolled between 1 and 10 patients, and 1,500 (55.1%) enrolled more than 10 patients. Registered sites encompassed nearly all hospital referral regions in the US, **Figure 2**. Sites participation occurred both in metropolitan and non-metropolitan areas and from different hospital types ranging from community hospitals to major teaching hospitals, **Table 1 and Supplemental Table 2**.

**Figure 1.**
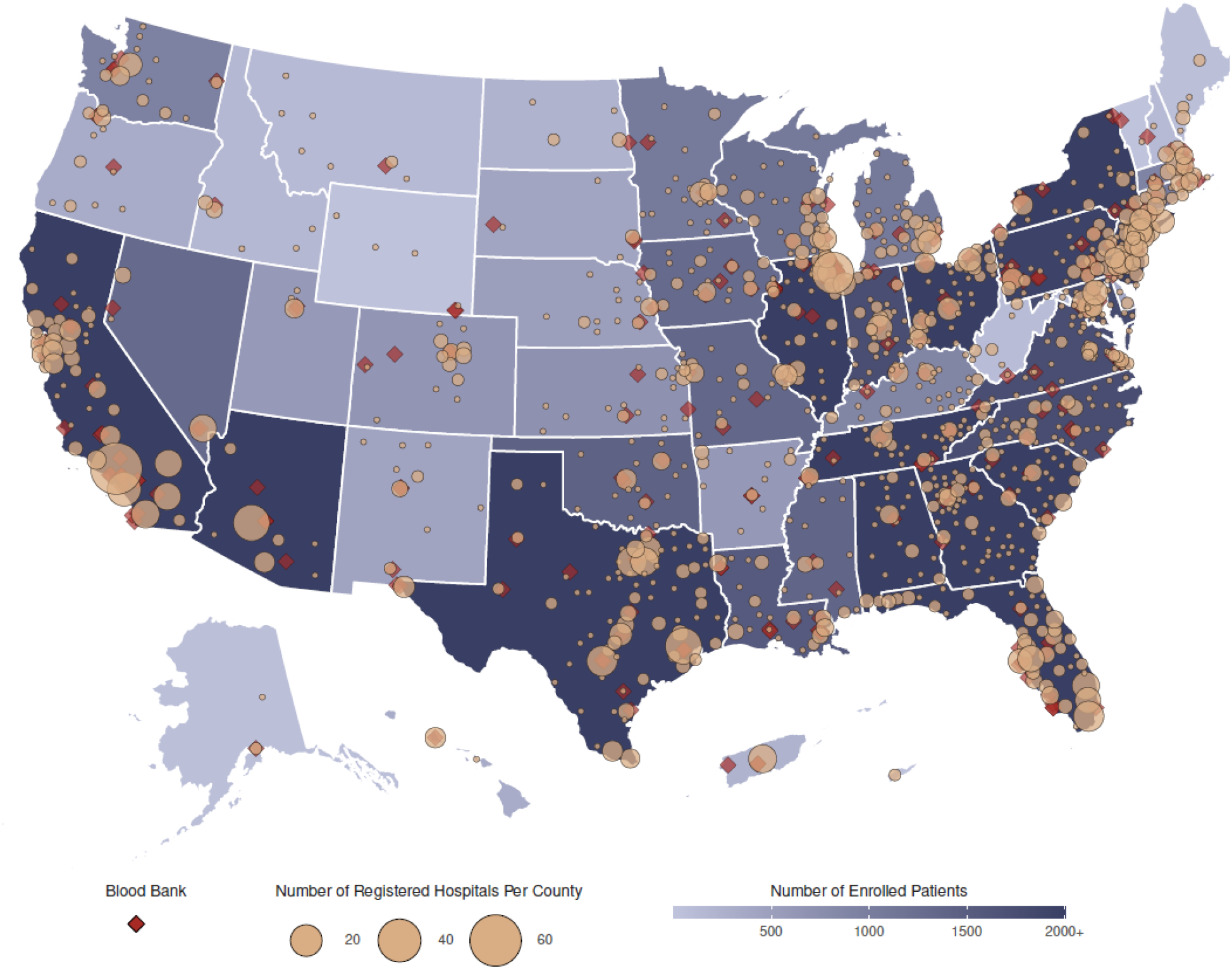
Participation in the US Expanded Access Program (EAP) to convalescent plasma. Choropleth map displaying the number of cumulatively enrolled patients in the EAP within each state of the US and participating territories, with lower enrollment values displayed in a lighter shade and higher enrollment values displayed in a darker shade of blue. Registered acute care facilities are represented as filled yellow circles, with larger circles indicating greater number of registered facilities within the county. Blood collection centers are represented as filled red diamonds. All sites with registered patients were included. The choropleth map does not display Guam or the Northern Mariana Islands.

**Figure 2.**
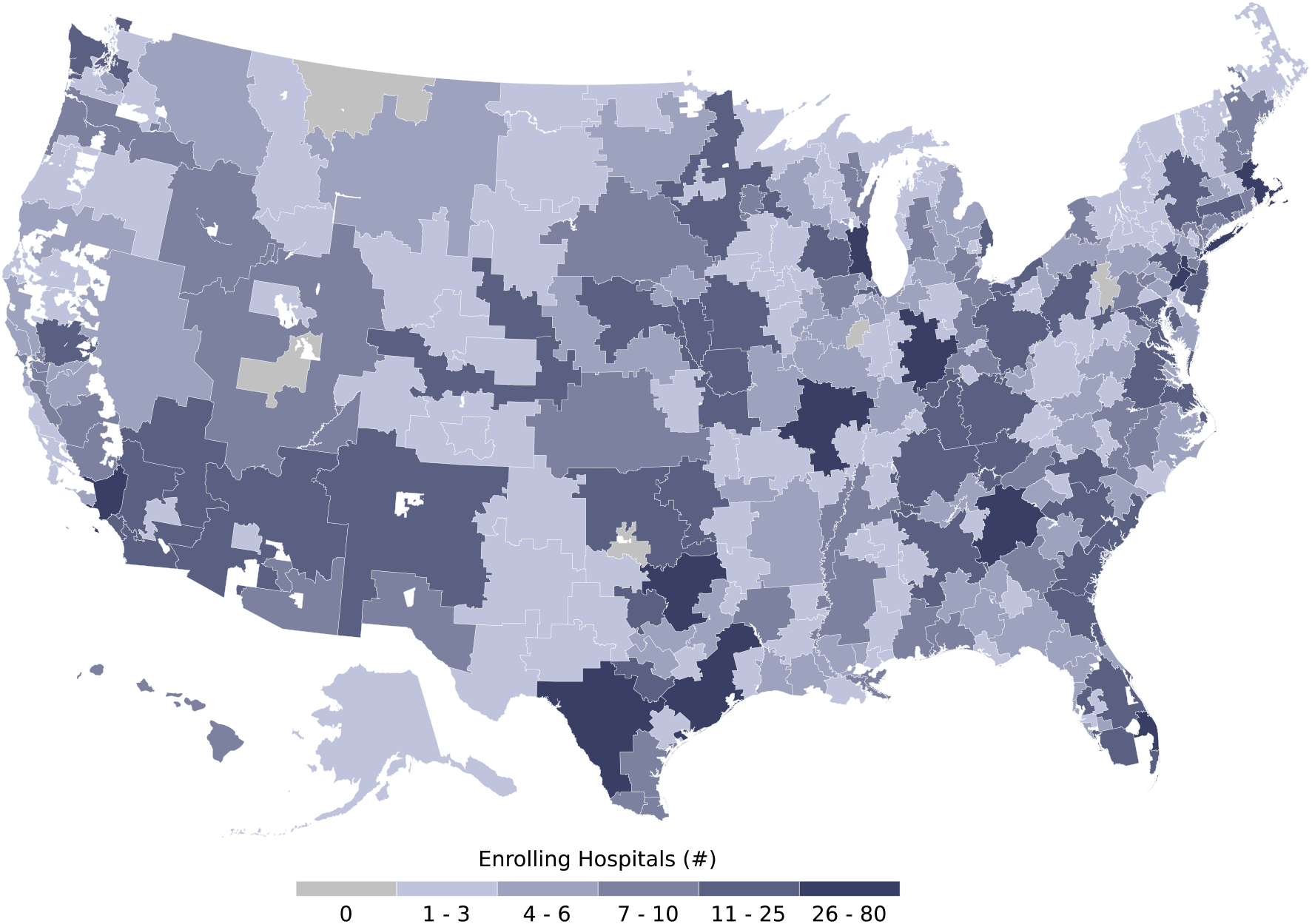
Participation of acute care facilities in the US Expanded Access Program (EAP) to convalescent plasma stratified by US hospital referral region. Choropleth map displaying the number of participating acute care facilities which enrolled patients in the EAP within each hospital referral region— a geographical regions which represent a catchment region of patients who get health care at similar facilities. Lower numbers of participating acute care facilities are displayed in a lighter hue of blue and higher numbers of participating acute care facilities are displayed in a darker hue of blue. Hospital referral regions with zero participating acute care facilities are displayed in grey. Hospital referral regions are not defined in US territories; thus, the choropleth map does not display data from Puerto Rico, the US Virgin Islands, Guam or the Northern Mariana Islands.

Recognizing the disproportionate effects of the COVID-19 pandemic on minority communities [17], the EAP sought widespread access to convalescent plasma including in all representations populations. Patient enrollment stratified by race and ethnicity groups is displayed in **Figure 3**, relative to 100,000 people from US census.

**Figure 3.**
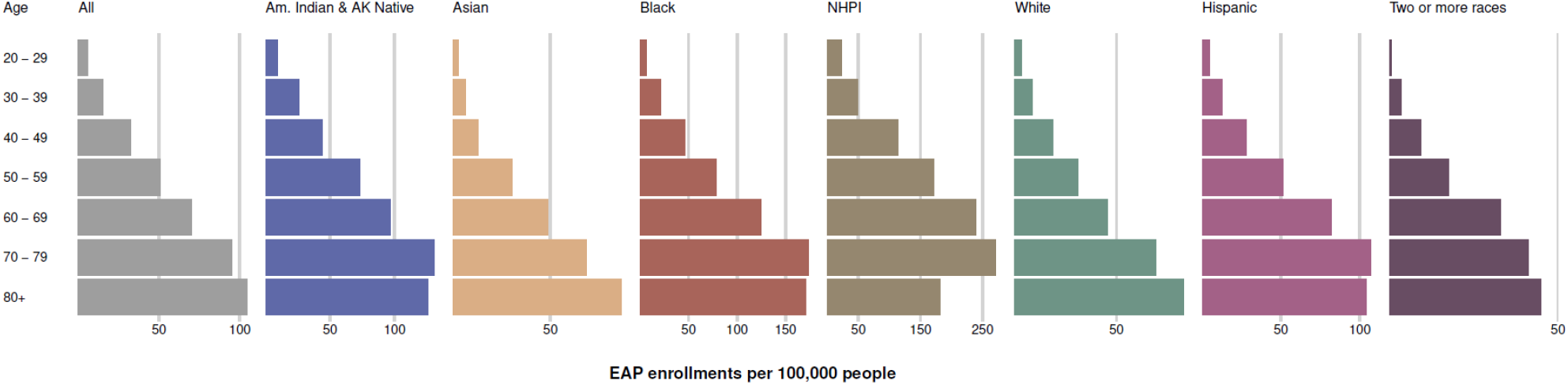
Patient enrollment in the US Expanded Access Program (EAP) stratified by race and ethnic group, relative to 100,000 people from US census per age, race and ethnicity. The length of each colored bar is proportional to the number of patients enrolled in the US EAP within the identified age group (years) and race or ethnicity category. The patient enrollment values are presented relative to analogous categorical data retrieved from the US Census Bureau.

### Temporal Trends in Enrollment

Enrollment in the EAP for convalescent plasma per US state on each day of the EAP is displayed in **Figure 4**. The individual state and regional aggregates show clear trends of when COVID-19 was surging during EAP enrollment. **Figure 5** presents enrollment over time together with the number of active COVID-19 cases per US state. Chronologically, increases in enrollment in the EAP closely followed the individual state infection rate. Enrollment in the EAP per 1,000 confirmed COVID-19 cases varied from 0.8 (Vermont) to 39.1 (Hawaii) patients across US states. The proportion that each US region contributed to the total enrollment into the EAP varied throughout the program and **Figure 6** displays proportional enrollment into the EAP across time by US geographical region, COVID-19 disease severity, and mechanical ventilation status prior to COVID-19 convalescent plasma transfusion.

**Figure 4.**
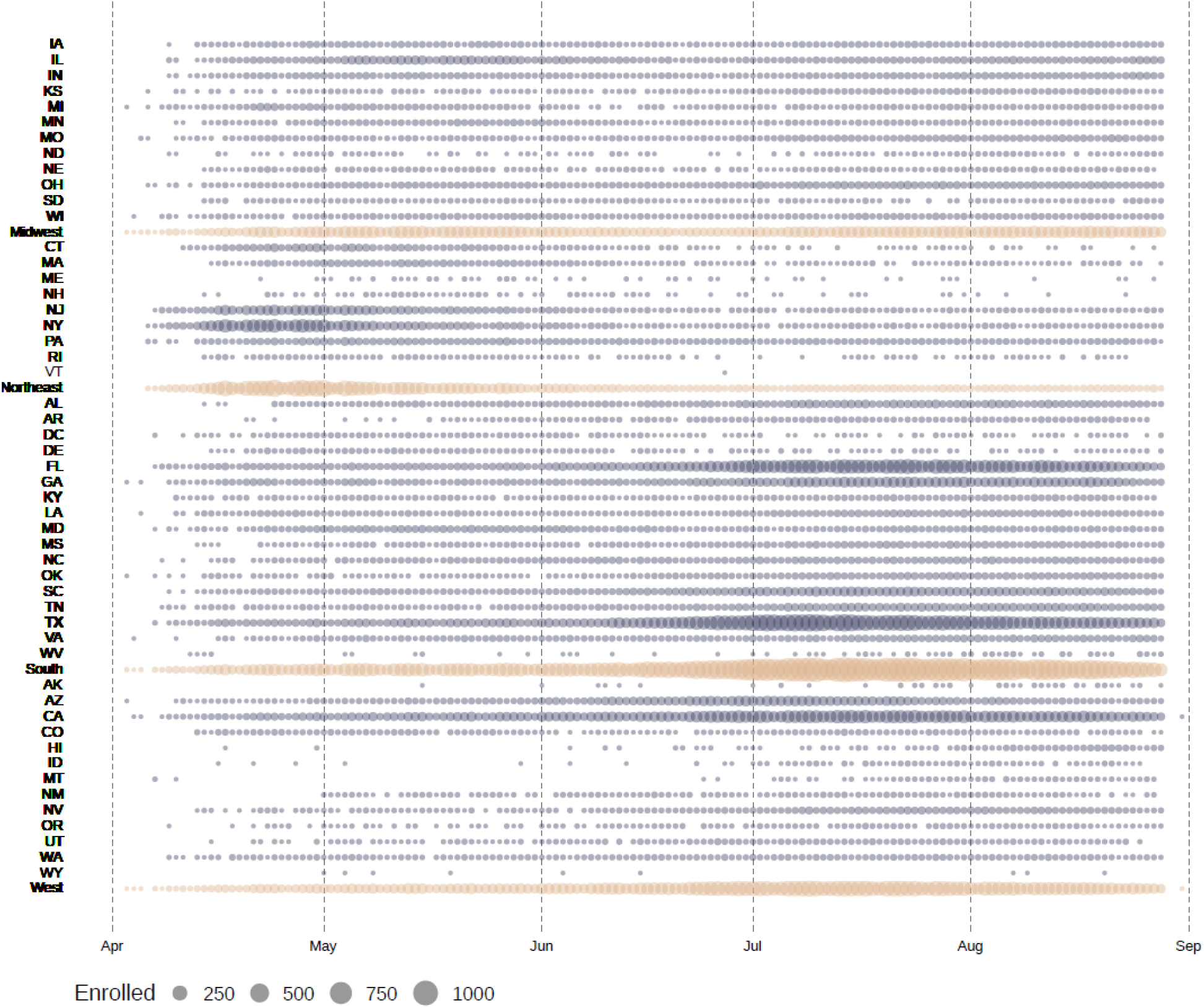
Daily patient enrollment in the US Expanded Access Program (EAP). Each circle represents one day in which at least one patient was enrolled, within the indicated US state or region. Blue circles represent daily US state enrollments and yellow circles represent daily US region enrollments. Larger circles represent a greater number of daily enrollments within the indicated US state or region. Nil patient enrollments on a given day are not represented with a symbol. States are ordered alphabetically by US region, and an aggregate for each region is also represented.

**Figure 5.**
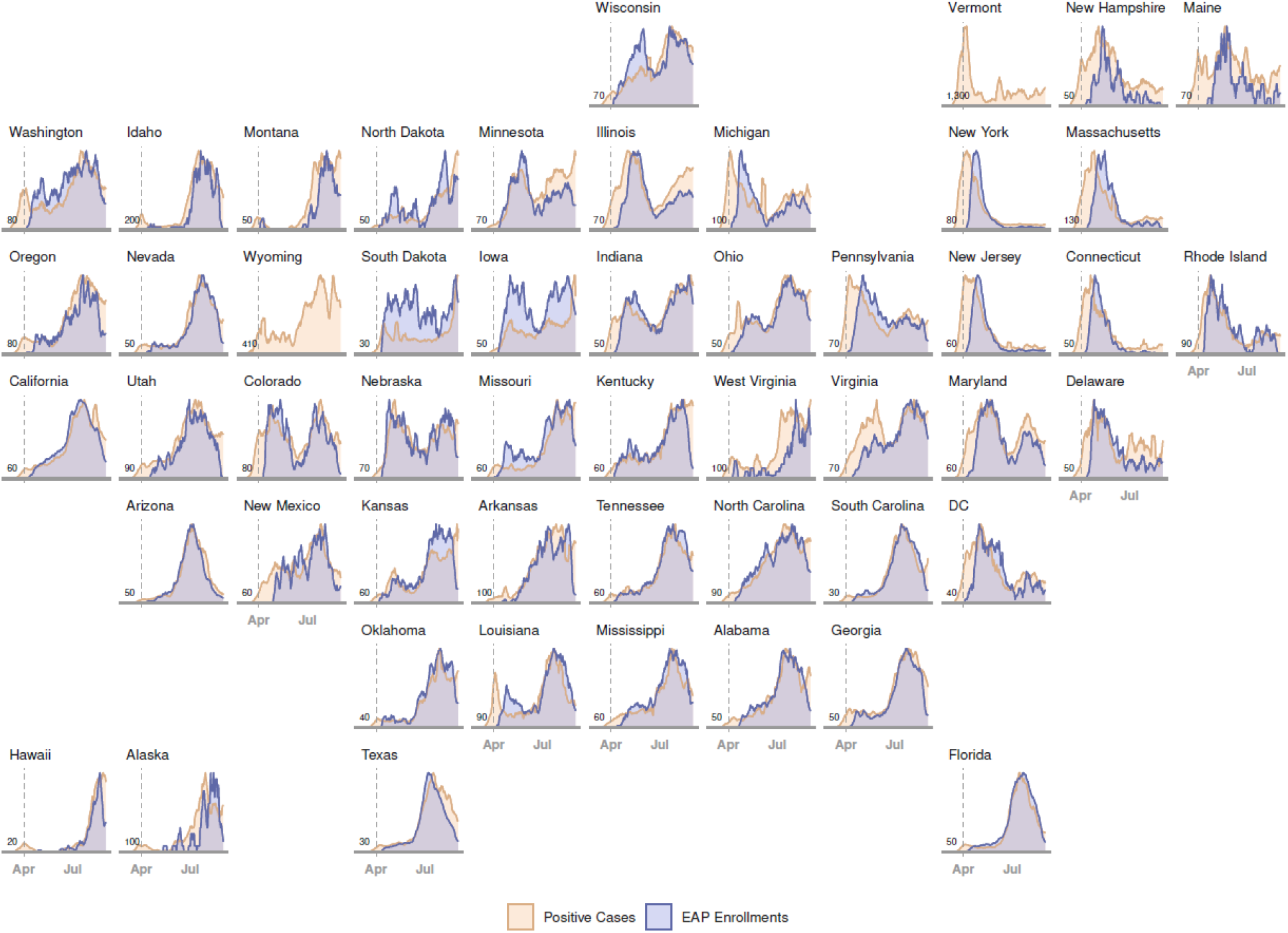
Daily rates of COVID-19-confirmed infections and patient enrollment in the US Expanded Access Program (EAP). Chronological line charts represent the daily number of state-wide COVID-19-confirmed infections and EAP patient enrollment sequentially arranged in a geofaceted depiction of the US. Daily rates are presented as a moving average across seven days, scaled between 0 (least cases/enrollments) and 1 (most cases/enrollments) for any day in each state. Vertical, dashed grey lines represent the start date of the EAP (April 3rd, 2020). Values in the lower left corner of each panel indicate the scaling factor between the two plots (cases/enrollments), which approximates the number of COVID-19 cases that contributed to one enrollment in the EAP. EAP enrollment data are not presented for Vermont or Wyoming because total enrollments were not greater than 10 patients.

**Figure 6.**
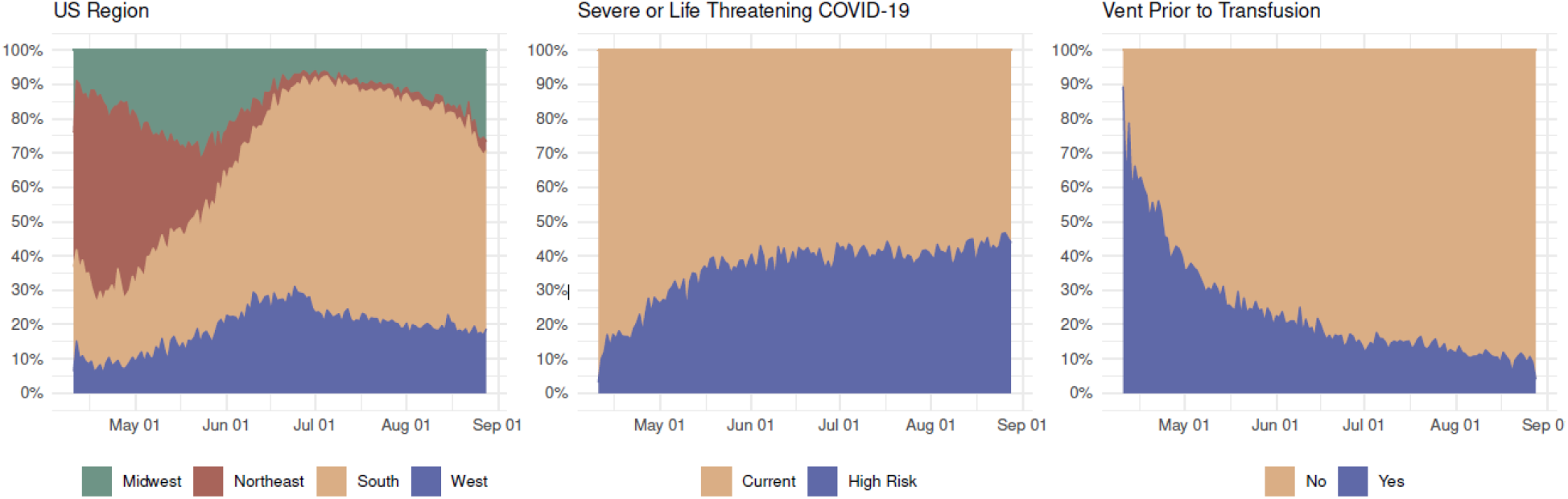
Daily patient enrollment in the US Expanded Access Program (EAP) relative to each US geographical region. Stacked area chart displaying daily rates of patient enrollment in the EAP as a proportion of the sum total daily enrollment within each of four US geographical regions, two categories of COVID-19 disease severity (severe or life-threatening), and dichotomous representation of mechanical ventilation status prior to COVID-19 convalescent plasma transfusion. Only patients that received a COVID-19 convalescent plasma transfusion are included in the rightmost panel dichotomized by mechanical ventilation status.

## Discussion

### Principal findings

The US EAP successfully provided access to COVID-19 convalescent plasma to over 105,000 patients, of which nearly 95,000 patients were transfused with convalescent plasma over the course of five months. Access to convalescent plasma closely kept pace with increases in confirmed US COVID-19 infections per state over time and there was substantial inclusion of vulnerable racial and ethnic minority populations. Geographically, enrollment in the EAP occurred in all US States, the District of Columbia, and the US territories of Puerto Rico, and the U.S. Virgin Islands. Patients were enrolled from all but five of US national hospital referral regions, and substantial enrollment occurred in both metropolitan and non-metropolitan areas.

### Demographic characteristics of patients enrolled in the EAP

Demographic characteristics of the complete enrolled EAP cohort showed substantial enrollment among patients over the age of 60 (57.8%), male patients (58.4%), patients of African American or Black race (18.2%), and patients of Hispanic or Latino ethnicity (37.2%). African American or Black race and Hispanic or Latino ethnicity make up 13.4% and 18.5% of the US population, respectively [27]. Previous studies have found that older [34, 35], male [35], African American or Black race [34, 36, 37], and Hispanic or Latino [37] individuals are at higher risk of hospitalization for severe or life-threatening COVID-19. The presented enrollment results from the EAP highlight that this program was able to provide access to COVID-19 convalescent plasma to demographic groups that have suffered the largest disease burden from the US COVID-19 epidemic.

### Chronological and Geographical characteristics

Peak enrollment in the EAP per day closely tracked the number of cases reported by state as the COVID-19 epidemic developed. Enrollment during April and May, 2020 was high in Northeastern states, whereas in July and August enrollment peaked in the Southeast and Southwest regions of mainland US, in line with the development of ‘hotspots’ in these regions over time [38]. An increase in enrollment in Midwestern states was observed during August 2020, and this trend was also closely associated with an increase in confirmed COVID-19 cases in that region. Although there was widespread participation across US states and territories, there were less than 10 patients enrolled in both Vermont (n = 1) and Wyoming (n=9), representing a small fraction of the total COVID-19 cases in those states. Given that there were no registered clinical trials involving COVID-19 convalescent plasma in these two states during the time of the EAP, there may have been regulatory or administrative barriers to participation in trials involving experimental therapeutics for COVID-19.

### Implications for future pandemics

The success of the EAP in providing rapid access to convalescent plasma in metropolitan and regional areas of the US that might not otherwise have had access to therapy provides a framework for future efforts when broad access to a treatment is needed in response to an infectious disease outbreak. In this regard it is noteworthy that given that clinical trials were limited to only a few institutions, that most patients treated with plasma would have had no access without the EAP or single-patient eINDs. The high use of convalescent plasma within the EAP indicates a high level of acceptance for this therapy by patients and frontline physicians despite the absence of high-quality data for its clinical efficacy. The EAP design was particularly effective in providing access to a potentially effective treatment in minority demographic groups and rural areas that are often underrepresented in US randomized controlled trials [39, 40].

### Methodological considerations

The US EAP for COVID-19 convalescent plasma aimed to provide access to a treatment possibly providing benefit, while randomized clinical trials were in various stages of development and enrollment. This enrollment report focused on the primary aim of the EAP of providing access to COVID-19 convalescent plasma; whereas, analyses on potential efficacy from EAP data have been presented elsewhere [19, 41].

### Limitations

Numerous challenges were encountered during the development and implementation of this national registry. Given the constraints on health care resources during the COVID-19 pandemic [42], this national registry used a modern design with creative solutions to overcome the epidemiological and contextual challenges of the pandemic [43]. These creative solutions included a central academic IRB for oversight, streamlined registration for sites and physicians, simple online data collection forms, robust support center accessible via email or telephone, limited patient exclusion criteria, limited restrictions on concomitant therapies, and no initiation or monitoring site visits. Several important limitations resulted from this design, however, including adjustments in required data collection elements which were inversely related to the number of cases of COVID-19 in the US, unavailable data due to abridged data collection forms, and missing data due to the nature of a national registry. Additionally, the EAP was designed to provide access to convalescent plasma at hospitals and acute care facilities that were not already part of a clinical trial or did not have the infrastructure to support complex clinical trials. This registry also did not require training of the local investigators or study team members. The design of this national registry provided widespread access to convalescent plasma and easy to complete data collection forms during a worldwide pandemic. This pragmatic approach did not ensure the highest quality of data nor completeness of data.

### Conclusions

The EAP provided rapid and broad access to convalescent plasma throughout the US and some US territories and was effective at providing therapy for demographic groups that were severely affected by COVID-19. Over time, the EAP provided access to convalescent plasma in response to sudden and exponential changes in SARS-CoV-2 infection rates. Data gathered from the EAP established the COVID-19 convalescent plasma was generally safe [20, 21] and provided key efficacy data that were an important component of the scientific evidence considered by the US FDA in the decision to issue an EUA [19] for convalescent plasma in the treatment of hospitalized adults with COVID-19. Hence, this program established that it is possible to obtain relevant and actionable safety and efficacy data during pandemic conditions. The efficient study design of the EAP may serve as an example for future efforts when broad access to a treatment is needed in response to a rapidly developing infectious disease providing access in areas typically underrepresented in clinical studies and thereby allowing capture of demographic groups that are often poorly represented in clinical trials.

## Data Availability

Individual participant data underlying the results reported in this publication, along with a data dictionary, will be made available to approved investigators for secondary analyses following the completion of the objectives of the United States Expanded Access Program to COVID-19 convalescent plasma. Limited and de-identified data sets will be deposited into a research data repository and may be shared with investigators under controlled access procedures as approved by the Mayo Clinic Institutional Review Board. A scientific committee will review requests for the conduct of protocols approved or determined to be exempt by an Institutional Review Board. Requestors will be required to sign a data use agreement. Data sharing must be compliant with all applicable Mayo Clinic policies.

## Article Information

### Disclaimer

The views and opinions expressed in this manuscript are those of the authors and do not reflect the official policy or position of the US Department of Health and Human Services and its agencies including the Biomedical Research and Development Authority and the Food and Drug Administration, as well as any agency of the U.S. government. Assumptions made within and interpretations from the analysis are not reflective of the position of any US government entity.

### Author Contributions

Drs. Joyner and Carter had full access to all the data in the study and take responsibility for the integrity of the data and the accuracy of the data analysis.

---

**Supplemental Table 1.**
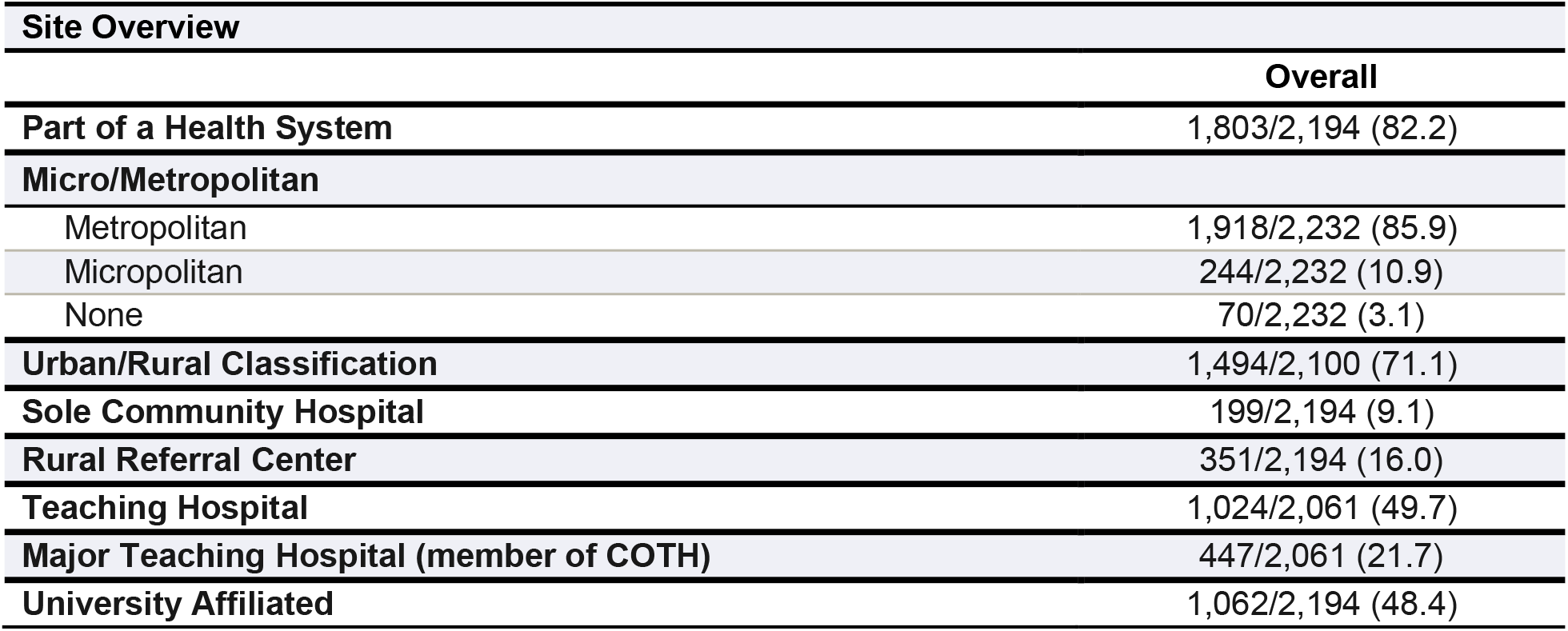
Characteristics of acute care facilities and hospitals that enrolled patients with COVID-19 in the US Expanded Access Program to convalescent plasma.

**Supplemental Table 2.** Pre-existing conditions and cardiovascular and lung disease history of patients with COVID-19 who enrolled in the US Expanded Access Program to convalescent plasma.

| Pre-Existing Conditions |  |
| --- | --- |
|  | Overall |
| **Pre-Existing Conditions** |  |
| History of lung disease (ex. COPD, lung cancer) | 7,225/44,081 (16.4) |
| Cancer other than above | 2,068/44,081 (4.7) |
| History of cardiovascular conditions | 21,673/44,081 (49.2) |
| Obesity | 14,608/44,081 (33.1) |
| HIV positive | 351/44,081 (0.8) |
| HCV positive (Hepatitis C) | 375/44,081 (0.9) |
| On immunosuppressive therapy | 1,631/44,081 (3.7) |
| Diabetes | 17,612/44,081 (40.0) |
| **History of Cardiovascular Disease** |  |
| Stroke | 1,493/19,490 (7.7) |
| Coronary Artery Disease (including prior myocardial infarct or MI) | 4,019/19,490 (20.6) |
| Valvular Heart Disease | 501/19,490 (2.6) |
| Peripheral Vascular Disease | 702/19,490 (3.6) |
| Hypertension | 16,894/19,490 (86.7) |
| Arrhythmias | 2,199/19,490 (11.3) |
| Heart Failure | 2,413/19,490 (12.4) |
| Myocarditis | 15/19,490 (0.1) |
| Cardiomyopathy | 586/19,490 (3.0) |
| Prior Coronary Artery Bypass Graft (CABG) | 688/19,490 (3.5) |
| Other | 1,959/19,490 (10.1) |
| None | 130/19,490 (0.7) |
| **History of Lung Disease** |  |
| COPD (chronic bronchitis, emphysema) | 2,403/21,193 (11.3) |
| Lung cancer | 160/21,193 (0.8) |
| Asthma | 1,301/21,193 (6.1) |
| None | 112/21,193 (0.5) |
| Other lung-related conditions | 744/21,193 (3.5) |

